# Beyond the numbers: A phenomenological analysis of women’s childbirth experiences in Spain’s evolving healthcare system

**DOI:** 10.1101/2024.06.13.24308819

**Authors:** Roser Palau-Costafreda, Sonia Nar-Devi, Maria Gil-Poisa, Amaia Pajares Manso, Ana España Vela, Noemí Obregón Gutiérrez, Ramon Escuriet, Mireia Julià, Ariadna Graells-Sans

**Author notes:** **Corresponding author:** Roser Palau-Costafreda, Escola Superior d’Infermeria Hospital del Mar. C/ del Dr. Aiguader, 80, Ciutat Vella, 08003 Barcelona. Twitter: @roser_palau.

## Abstract

**Background:** Childbirth is a transformative experience, yet a considerable percentage of women worldwide encounter negative birth events, affecting maternal wellbeing and mental health. The choice of birth setting significantly impacts outcomes, with midwifery-led units often associated with lower intervention rates and higher satisfaction levels. The recent introduction of midwifery-led units in Spain presents a unique opportunity to explore the impact of this model within a medicalized healthcare context.

**Aim:** To capture the depth and diversity of women’s voices, understanding factors influencing their perceptions of childbirth experiences following the introduction of the first midwifery-led unit in the Spanish Healthcare System.

**Methods:** A qualitative study with a phenomenological approach within the constructivist paradigm. Four focus groups were conducted including nineteen women who gave birth in a hospital with both an obstetric and a midwifery-led unit.

**Findings:** Three main themes were identified; ‘Shaping birth expectations’, highlighting the influence of cultural and social contexts on women’s childbirth expectations; ‘The childbirth essentials’, incorporating fundamental characteristics related to the model of care; and ‘Navigating the protective factors’, considering the pivotal role of midwives in delivering compassionate and respectful care.

**Conclusion:** These findings offer valuable insights into childbirth experiences, advocating for a transformation of the medicalized healthcare system in Spain through the integration of midwifery-led units. By prioritising women’s voices and addressing systemic inequalities, healthcare policymakers can enhance maternal care practices and foster positive childbirth experiences for all women.

## Introduction

Childbirth is a deeply personal and transformative experience encompassing physical, emotional, and social dimensions that shape the identity and memories of those involved^1^. The significance of childbirth as a multifaceted experience resonates globally, influencing the well-being of both women and society at large. The World Health Organization (WHO) underscores the importance of accessible, acceptable, and high-quality healthcare^2^, emphasising a ‘positive child experience’ as a crucial outcome indicator for women undergoing childbirth^3^. A comprehensive definition of a positive childbirth experience emphasises feeling supported, in control, safe, and respected, highlighting the role of provider interactions^4^. Positive childbirth experiences not only contribute to women’s psychosocial well-being, but also hold the potential to empower and transform individuals^5^.

Despite global efforts to prioritise respectful maternity care^6^, evidence suggests that a substantial percentage of women worldwide experience childbirth as a negative or traumatic event, with prevalence ranging from 4% to 45%^7, 8^. The repercussions of such negative birth experiences are profound, closely linked to disrupted maternal psychological and emotional outcomes, including postpartum anxiety, post-traumatic stress disorder and postpartum depression^7, 9, 10^. These consequences extend to maternal self-esteem, the ability to bond with the infant, breastfeeding rates, and the overall transition to motherhood^11, 12^.

The choice of a birth setting significantly influences a woman’s birth experience^13^. While the majority of births in high and middle-income countries occur in a hospital, research indicates that midwifery-led units (MLUs) or birth centres, are associated with reduced rates of medical interventions during labour and childbirth when compared to traditional obstetric units (OU)^14,15^ and higher satisfaction rates^13^, attributed to feelings of comfort, active involvement, and increased sense of control^14,16^. These findings indicate the importance of considering diverse birth settings when evaluating the overall experience of childbirth.

Spain’s approach to childbirth care is undergoing a transformative shift through the implementation of MLUs in its National Health System^17, 18^. This approach presents a unique opportunity to explore the impact of this on women’s birth experiences in Spain, given the country’s prevailing medicalized healthcare environment^19^. Despite recognizing the significance of factors like continuity of care, midwifery-led models, patient-centred care, provision of information, and participation from decision-making^13, 14, 20^, these elements have not been fully integrated across the Spanish Healthcare System, leaving the complexity of childbirth care in Spain only partially understood. Exploring professional practices and various factors influencing birth experiences within this context will contribute valuable insights, advancing our understanding of the implications of this model for maternal well-being.

The aim of this study is to capture the depth and diversity of women’s voices, understanding the interplay between the factors influencing their perceptions of the childbirth experience following the introduction of the first MLUs in the Spanish Healthcare System. The findings aim to provide useful information to healthcare providers and policymakers to promote positive childbirth experiences.

## Methods

### Design

This study employs a qualitative methodology and the research design is situated within the constructivist paradigm, offering an ontological approach rooted in the understanding of realities through situated and subjective construction. Phenomenology has been selected as a theoretical and methodological framework facilitating a deep exploration of childbirth experiences within their socio-cultural context^21^. The philosophical alignment of phenomenology with midwifery^22^ accentuates key concepts including care, presence, concern, interpersonal dynamics and therapeutic caring connections.

### Setting

This study was conducted in a community public Hospital in XX, Spain, with an average of 650 childbirths annually. The hospital offers two distinctive birthing settings: a traditional OU and a MLU. Although the MLU is separate from the OU, it has access to the same hospital facilities.

The OU follows a biomedical model of care, staffed by obstetricians, anaesthetists, paediatricians, and midwives. It is equipped to handle high-risk pregnancies and complications during labour and birth. The OU features advanced medical equipment, including emergency surgical facilities. Pain management options such as epidurals are readily available. The care approach in the OU involves standardised medical protocols and interventions, including inductions, caesarean sections, and assisted births.

In contrast, the MLU, introduced in December 2017 as the first of its kind in the Spanish Healthcare System, is managed exclusively by qualified midwives and operates autonomously within the hospital. The MLU focuses on women with uncomplicated pregnancies and aims to provide a home-like, calming environment. Medical interventions are limited, with an emphasis on physiological birthing practices. Facilities in the MLU include a birthing pool and other natural pain relief methods. The MLU follows a biopsychosocial model of care with a women-centred approach, featuring individualised care plans that encourage active participation of the mother in the birthing process.

### Participants and sampling

Purposive sampling was employed to ensure diverse participant backgrounds. Inclusion criteria were: (1) Spanish-speaking women; (2) residing in Spain; (3) aged 18 years or older; (4) with uncomplicated pregnancies; (5) who had given birth either in the MLU or the OU. Exclusion criteria comprised: (1) births occurring before 37 weeks; or (2) high-risk pregnancies.

Participants who were already part of the BirthingBetter study were identified and subsequently invited to participate in this research by a member of the research team via telephone. Following this initial contact, detailed information and informed consent forms were sent to them via email. Participants were given time to consider their involvement before being invited to join one of the four focus groups, scheduled between 4 and 9 months postpartum.

Out of 25 women that were invited to the focus groups, 22 consented to participate, and three withdrew last-minute changes in family arrangements. Comprehensive data, including background information and birth details, were systematically collected through online questionnaires. The characteristics of the participants are summarised in Table 1.

### Data collection

Data were collected through focus groups, selected for their capacity to create rich, interactive discussions. The focus groups were conducted to resemble postnatal or breastfeeding support groups, reflecting community practices where women openly share their childbirth experiences. Focus groups were preferred over individual interviews to provide a comfortable, supportive environment, enhancing participants’ willingness to share personal and sensitive experiences. This method is particularly effective in uncovering cultural norms and collective attitudes toward childbirth that might not surface in individual interviews.

Nineteen women participated in four focus groups conducted between March and April 2022. Each session was facilitated by a midwife and observed by an additional researcher who took field notes. To accommodate postpartum women and minimise travel, sessions were held virtually via Zoom^23^. Participants consented to audio and video recording for transcription. The sessions, conducted in Spanish and Catalan, lasted an average of 80 minutes, ranging from 68 to 102 minutes. The interview guide (Table 2), developed based on literature and input from women and healthcare professionals, included five open-ended questions exploring childbirth experiences and related factors. Transcripts were initially in Spanish and then translated into English. Any real names from mothers or healthcare professionals were fully replaced with pseudonyms. Once data saturation was achieved during the focus groups, no further groups were conducted, ensuring the data’s depth and richness were sufficient.

### Data analysis

Following the collection and verbatim transcription of all narratives, a thematic analysis was conducted, drawing on established methodologies^24^ and using the Atlas.ti program. To ensure the robustness and precision of interpretation, three members of the research team (XX, XX, and XX) initially performed independent analyses. The accuracy of the code grouping process underwent validation through discussions involving the other researchers, resulting in a consensus on the categorization of codes into themes and subthemes, also minimising biases linked with the professional background. This collaborative approach allowed for the triangulation of information and results, enhancing the reliability of the findings. Themes were derived from the data, and any discrepancies that arose were resolved through consensus among the team members. In the final phase of analysis, three overarching themes with their subthemes were formulated and are presented in Table 3.

### Ethical and quality considerations

Ethical approval (registration number XX) was obtained from the Ethics Committee of XX. Anonymity protection was prioritised due to the study’s sensitive nature and small sample size. Data was pseudonymized and securely stored by the lead researcher, in accordance with the current data privacy regulations. Participants received a participant information sheet detailing the study and were given a two-week period to decide on participation. They signed a consent form to confirm their voluntary involvement. Participants were assured the right to withdraw without consequence, even during focus group discussions, ensuring their decisions were well-informed and voluntary.

## 3. Results

The childbirth experiences of women were grouped into three themes: ‘Shaping birth expectations’, ‘The childbirth essentials’, and ‘Navigating the protective factors’. Within these 3 main themes, 10 subthemes were identified (Table 3).

### Shaping birth expectations

#### Cultural and social factors

Participants commonly recognized the profound impact of childbirth expectations on the overall birthing experience. The childbirth experience was significantly influenced by the alignment or misalignment of these expectations, which could determine whether the experience was viewed positively or negatively. The shaping of these expectations appeared deeply rooted in cultural and social contexts. Women particularly emphasised the role of external influences in shaping their expectations, attributing this phenomenon to the diminishing visibility of authentic birthing experiences within the community. For example, relying on external sources such videos and peer discussions to form their childbirth expectations:

> *‘I’ve read all the birth stories, watched all the videos. […] I would ask my friends about it. I like it. But, in real life, we don’t have that many chances. I haven’t seen a birth yet, you know? I have never seen a vaginal birth more than the ones that I’ve seen on YouTube. I mean, it’s something we talk about a lot, that we prepare a lot for, but, in reality, until you find yourself in that situation, you’ve never been present in one.’* (P2, OU)

Moreover, the societal association of natural childbirth with strength placed significant pressure on women, compelling them to exhibit resilience and endurance during labour. Participants frequently expressed feelings of guilt, perceiving themselves as having ’failed’ if they deviated from societal expectations of a natural birth. Participant 15’s narrative illustrates this internal struggle, as she initially aspired to a natural birth in the MLU but ultimately underwent labour induction in the OU:

> *‘And always, when I think about it, I sometimes feel a bit guilty. Sometimes I think ’what if,’ ’what if I had done it,’ ’what if…’. Well, all those thoughts, right? that mothers always have… I don’t know. I panicked, and it was like I couldn’t handle it, you know? I was already worn out, and no, I couldn’t… I couldn’t.’* (P15, OU)

### Previous obstetric experiences

For some participants, a pivotal moment was linked to giving birth for the first time. The birth of their first child became transformative, shaping expectations for subsequent pregnancies, particularly those marked by dissatisfaction. The proactive approach stemmed from a strong desire to avoid reliving challenging memories associated with previous childbirths:

> *‘I was coming from an experience that was not respectful with my first daughter. I did the birth plan, everything, we prepared everything with my partner, and well, that day it just stayed there on a shelf. […] Everything I didn’t want to happen, it happened. I had bad memories… and my intention was not to have more children. But this time I wanted to have a good memory, and that’s why I came [to the MLU], and… and it was very, very beautiful and respectful.’* (P7, MLU)

### Knowledge, interaction and healthcare accessibility

The accessibility and comprehension of the healthcare system emerged as another crucial determinant in shaping childbirth expectations and influencing decisions regarding birthing environments. One participant referred to the unequal information regarding the options for place of birth:

> *‘Information is something that we should have from early in pregnancy. Because, even when you go for check-ups, unless you don’t actively seek information, you might remain unaware of places like this [MLU] and alternative ways of giving birth.’* (P9, MLU)

Furthermore, the ongoing interaction with the healthcare system played a significant role in shaping expectations. For example, external factors, including medical opinions, seemed to contribute to the gradual formation of mindsets and childbirth expectations:

> ‘*My girl was born big, well, 3.8 kilograms. There are much bigger babies. But, from the very beginning, in the community centre, they told me I was having a very big girl and that I wouldn’t be able to deliver naturally. There was no support anywhere. […] They gradually undermined our confidence, and although we appeared strong externally, at every medical check-up, we found words or phrases that didn’t sit well with us.’* (P10, OU)

### The childbirth essentials

Women consistently articulated the essential elements that contribute to enhancing their overall satisfaction. In this theme, we describe these elements within the two distinct models of care—the MLU and the OU.

### One to one care

In the MLU, participants usually recounted experiences of one-to-one care provided by a dedicated midwife during labour. This personalised attention significantly contributed to a sense of support and individualised care throughout the birthing process:

> *‘Raga, the midwife, was there accompanying me and supporting me at all times. And it was somewhat thanks to her that I was able to have a birth that I would have never imagined, it was really beautiful. […] The last few hours were very intense, but at the same time, I felt very supported at all times. I remember a moment that was really cool, you know? It was during the shift change of the midwives, but Raga, who had been with me throughout, didn’t want to leave me alone, you know? So the midwives were there, and it was the moment of delivery, and they were all encouraging me. […] I remember this moment as really beautiful, and everyone was there, waiting for the baby to be born. […] I felt very supported with the encouragement I really needed at all times.’* (P6, MLU)

Conversely, participants in the OU described instances where they felt a lack of support and a desire for more personalised attention. Participants expressed frustration with the workload of midwives, as the example of this participant:

> *‘And what if I could have done a bit of biomechanics to see if the baby had positioned well? But what happened? After getting the epidural, there was no one with me either. […] The midwives were extremely busy; there were three or four other moms arriving suddenly at midnight, and it was 1 am. I was there alone, waiting to see when my time would come.’* (P1, combined care)

### Pain relief strategies

Within the MLU, participants appreciated the holistic approach to pain relief, acknowledging the diverse needs of women during childbirth. The model’s commitment to addressing individualised pain relief needs was exemplified in Participant 17’s encounter with a midwife named Claire:

> *‘Claire, who was wonderful, […] seemed like Mary Poppins because she was pulling out all kinds of resources. She would come in and say, ’come on, why don’t we try the electrodes,’ ’why don’t you try the ball,’ ’why don’t you shower,’ ’why…’ And all the time like that, right? And I was drawing strength, I don’t know where from.’* (P17, combined care)

Conversely, in the OU, participants perceived a more limited availability of pain relief strategies, primarily centred around epidural-based options, with reported limitations in accessing alternative methods like showers, bathtubs, or birthing pools. Participant 13’s experience highlighted the perceived constraints in the model:

> *‘I couldn’t use the shower because another woman was using it, and I asked for a bathtub or the birthing pool. They told me it wasn’t possible in the labour ward.’* (P13, OU)

### Privacy and dignity

Concerning privacy and dignity, participants, emphasised the utmost significance of preserving dignity, privacy, and autonomy throughout the childbirth experience:

> *‘We were super well; we have very, very good memories. We were accompanied all day by Hannah [the midwife]. Since we entered the room, she set up the entire place as if we were at home. With soft light, salt lamp light, aromatherapy… I really valued having privacy. Especially when you have to be there for many hours. Being able to be calm, knowing that no one will enter the room, except for her [the midwife]. That also helped me a lot to be calm and to be able to be like home.’* (P8, Combined care)

The transition between rooms was noted by women like Participant 13, who articulated a moment of discomfort in the transitional zone. The disparity in ambiance was highlighted as important for maintaining a sense of privacy.

> *‘I went up to the room [antenatal ward], and they told me that when the contractions were continuous and strong, I should let them know, and I could come down [to the OU]. So, when I thought it was time I asked to come down. There was no dim lighting, I mean, it was a cold hospital room with bright white light that was blinding me. There were people walking up and around, and I felt a bit defenceless.’* (P13, OU)

Furthermore, Participant 16 reinforced the significance of ambiance in cultivating a sense of intimacy when talking about what was essential for her to have a positive birth experience:

> *‘I think that privacy is important. The privacy that you have, right? In the room, in the entire environment. I believe that for giving birth successfully you need this privacy… the low light, minimal noise. So, you can enter into your own world. I think that it is important.’* (P16, Combined care)

### Respectful care

There was absolute consensus among participants on the significance of respectful care on their childbirth experiences. This can be seen through the words of this participant, that expresses a deep appreciation for the respect received:

> *‘For me, the most crucial aspect was the respect I felt throughout, and the support. […] I felt like I was in harmony, right, with the midwives and such. Above all, I felt very respected. It really bothers me when they infantilize me; it makes me very angry. I cannot tolerate it. This feeling of ’I am the doctor, and I know, and you don’t’. That paternalistic attitude, ’I am big, and you are small,’ I can’t stand it. For me, this is the main aspect.’* (P15, combined care)

Reflecting on the limited emphasis on the woman’s experience and emotions within the OU, participants reported a sense of diminished respect and attention to their emotional well-being. The following participant vividly describes her experience upon going to theatre for a caesarean section:

> *‘As I entered the operating room, I was… It was a drama, feeling nauseous, trembling. In the operating room, no one said anything to me. Until at one point, I said, poking my head through the surgical screen, ’Hey, today is a very important day for me.’ I didn’t say it to seek attention at that moment, but because I wondered if someone would look into my eyes at any point. That really bothered me because it felt like me on one side, [of the surgical drape] and another world going on the other.’* (P2, OU)

### Navigating the protective factors

#### Meeting “elements of value” in care

During childbirth, despite facing unmet expectations, women emphasised “elements of value” that triggered positive emotions or acceptance of the experience. Participant 10 summarised several of these “elements of value”, encompassing patient-centred care, individualised attention, emotional support, accompaniment, and compassionate care, despite undergoing a caesarean section:

> *‘I ended up having a caesarean section, but I was very clear about the birth plan. I was able to discuss it calmly, and the medical team there was very pleased because they told me it was the first caesarean they had done with such respect. They lowered the curtain; I could see the baby coming out. We did skin-to-skin immediately. My husband was there; he could record everything. They turned off the lights, which they had never done before. I asked them to turn off the lights and leave only the illumination on the belly. Well, despite it not being what we wanted, it wasn’t so bad in that regard.*’ (P10, OU)

Participants highly valued the sense of control, manifested through various means, including keeping them informed and engaged during interventions, providing continuous updates on administered medication, explaining procedures, and detailing the childbirth process. For instance, this participant stated:

> *‘When they said, “we’ll need vacuum extraction”, I felt the only moment of panic. My legs were shaking uncontrollably. I vividly remember grabbing Shavonne’s [the midwife] hand tightly and telling her, “Shavonne, I need you to explain how this will go.” She showed me the vacuum and explained each step with detail. That’s when I started calming down.’* (P5, OU)

### Midwifery role on ‘childbirth essentials’ and ‘elements of value’

The provision of close, understanding, and humanised care, particularly by midwives, remains integral in shaping a positive birthing experience. Their presence and support, without overshadowing the woman, her chosen companion, and the new-born, significantly impacted the overall childbirth experience. Participant 8 exemplified this sentiment through her description of the connection and trust with Hannah, the midwife:

> *‘The connection I had with Hannah [the midwife] was… well, that atmosphere… well, she, yes, so familiar, so close… We were lucky, or not (laughs), that we got her a couple of times [on antenatal appointments]. And then, in those two times, we established a kind of relationship, and… and we already knew that she would be there on Monday, and since the baby had to be born on Monday no matter what, we already knew she would be there, and that gave me a lot of peace, you know? Knowing she would be there and knowing that Hannah would be super sweet, and well, that treatment, like super close and as if we had known each other all our lives.’* (P8, Combined care)

However, when instances of a lack of connection with midwives were reported, feelings of loneliness emerged:

> ‘*Some time has passed now, so I can talk about it more comfortably. I understand that everyone is different, and I was at the hospital for forty-eight hours, so I saw many people come and go, but with the midwife who came in the morning, I don’t know how to call it. Empathy? I felt a lack of presence, you know? I wasn’t asking her to be by my side all the time, but not absent either, I just felt that she wasn’t as present. And that started to affect me. […] I felt super alone.’* (P18, combined care)

### The role of advocacy for women’s empowerment

Advocacy was important for participants during childbirth, providing support even when decisions didn’t match their preferences. Women valued having someone, often a midwife, advocate for their expectations and desires:

> *‘I felt that she [the midwife] had read my birth plan, understood my desires, and even though there were decisions that I didn’t truly expect, I sensed that I had some influence. She became my voice in the team, advocating for what I wanted […] Always striving for that extra time and the least medicalized approach possible. She also listened to my partner, for instance, when he conveyed, ‘No, she wants it to be in the water.’ Throughout the day, she remained very attentive to both of us, always there if we needed anything, and ensuring privacy.’* (P8, combined care)

Women who felt more supported during childbirth described a more positive overall experience and a sense of empowerment. Feeling empowered, listened to, and supported during labour extended beyond the birthing process:

> *‘What I liked was that Hannah and Sofia [the midwives] accompanied me, and honestly, I really liked how they intervened. The way they… They were present [..] I felt like the protagonist, me and my baby. That was really beautiful. You feel that you are capable. They would say, ‘Yes, you can,’ and I don’t know… It’s a very beautiful support, really. And for me, having given birth there, well, the truth is that… I will have a very good feeling from when I had my baby until… well, throughout my life, right? Because I think childbirth… it marks you, right? As a woman. […] For me it was incredible and magnificent, and I will never forget it. For my little one, surely too, and well, for my family in general, for my partner, and for everyone, well… Fantastic… (laughs)’* (P9, MLU)

## Discussion

Our study delves into the complexities of women’s childbirth experiences within the evolving Spanish Healthcare System, with a focus on the newly introduced MLUs. Firstly, our findings highlight the central role of women’s expectations, deeply influenced by cultural norms, personal experiences and societal trends. Unmet expectations pose considerable challenges for most women^25^. Traditionally a communal event, changes in culture and medical practices have shifted childbirth towards a more private affair. This transformation has altered the visibility of real birthing experiences, introducing an element of uncertainty for expectant mothers. Our findings are in line with Lawrence, Richardson, and Philp^26^ where external sources like social media shape these expectations, often adding an additional layer of pressure on the societal impositions that associate natural childbirth with strength, contributing to feelings of guilt when the actual experience deviates from this expectation^27^. Previous childbirth experiences profoundly impact subsequent expectations, with dissatisfying experiences driving proactive shaping of future expectations, aligning with the concept of redemptive birth explored by Thompson and Downe^28^.

Importantly, our findings reveal disparities in accessing MLUs within the Spanish healthcare context. Uneven information presents challenges for women seeking an MLU, often relying on information from friends or personal research^29^. Variations in awareness not only shape individual childbirth perspectives but also influence expectations from the healthcare system and place of birth choice^30^, raising concerns about embedded inequalities, where expectations are inherently tied to what one can access and imagine. Comprehensive education, positive pregnancy experiences, and diverse birthing model accessibility are essential for targeted support, urging healthcare professionals and policymakers to address these multifaceted influences to mitigate inequalities and vulnerabilities in maternal care.

Secondly, the model of care, as well as the role of the midwife, serve as a mediator of the childbirth experience, acting as catalysts between expectations and the actual birthing process, influencing the overall perception. Here once again the perception of this role is culturally interpreted, although consensus is identified regarding the fundamental characteristics it should encompass: respect, dignity and autonomy. Women in our study highlight the advantages of a biopsychosocial approach characterised by patient-centeredness, informed choice, and holistic support^31^. The MLUs, inherently women-centred, align with the essential elements for a positive birthing experience outlined by Warren et al.^4^. Conversely, the OU, grounded in a biomedical model, often falls short in meeting these elements, emphasising the need for a shift towards more patient-centric models^32^. Furthermore, the systematic review by Downe et al.^16^ also highlights giving birth to a healthy baby in a safe environment as an important factor.

The role of midwives in delivering compassionate, understanding, and humanised care significantly influences the overall childbirth experience, resonating with the works of Dahlberg et al.^33^. Furthermore, trust and a sense of connection with the midwife during crucial moments were highlighted as important aspects, aligning with the concept of “watchful attendance,” proposed by de Jonge, Dahlen, and Downe^34^, underlying the midwife’s role in being fully present and attentive to individual needs, though challenging to quantify. This dynamic aligns with the concept of agency, where women perceiving control over the childbirth process, even when it deviates from expectations, report more positive birthing experiences^35^. Thus, the midwife’s role inherently involves acting as a health advocate, materialised through the preservation of the woman’s agency. This is consistent with the findings from Watson et al.^36^ and Kuipers et al.^37^. Feeling empowered, listened to, and supported during labour extends beyond the birthing process, influencing women’s identity as mothers and their broader life perspectives ^38^. Our study aligns with Michels, Kruske, and Thompson^39^, indicating that a positive perception of childbirth correlates with increased self-esteem, self-efficacy, independence, and empowerment. These findings underscore the far-reaching impact of midwifery care on women’s well-being, emphasising the need for continued efforts in fostering positive connections and support throughout the childbirth journey.

A key finding of this study is that elements inherent to the biopsychosocial model of care are transferable to the OU, acting as protective factors against a negative experience. Therefore, it seems that the establishment of MLUs in the Spanish Healthcare System facilitates the integration of these characteristics into the biomedical model, influencing OU practices. These findings highlight the importance of tailoring care models to individual preferences and needs, advocating for a patient-centred approach in maternity care. The call to move towards a more biopsychosocial model, while ensuring the safety of childbirth interventions, aligns with the necessity for an individualised approach and a broader shift towards a patient-centred paradigm in healthcare.

The primary strength of this study is its role as the first qualitative investigation into childbirth experiences post-implementation of the first MLU in the Spanish Healthcare System. However, qualitative data, although rich in narrative depth, may present challenges regarding generalizability and replicability. Secondly, the sample’s predominantly Western European, highly educated composition could limit the diversity of perspectives captured in the study. Additionally, another limitation was not identifying more variables of the socio-economic status of the participants, which could have provided a more comprehensive understanding of the varied childbirth experiences. While online interviews offer convenience during the postnatal period, in-person groups may be more fitting when addressing emotionally sensitive topics. Nonetheless, efforts were made during the focus groups discussions to foster a safe environment to encourage the expression of any type of feelings. Despite these limitations, the study’s adoption of a constructivist paradigm and a phenomenological approach ensures a thorough exploration of women’s lived experiences. Another strength of this study is the independent analysis of the data by researchers from different professional backgrounds, which helps to avoid biases and enrich the interpretation of the findings.

## Conclusion

Our research provides a comprehensive understanding of the factors influencing women’s childbirth experiences within the Spanish Healthcare System, particularly following the introduction of MLUs. Our findings highlight the significant influence of childbirth expectations on shaping childbirth experience, which are profoundly influenced by cultural, social, and healthcare contexts. Disparities in information access and MLUs availability reveal underlying inequities in childbirth decision-making, emphasising the necessity for inclusive education and easily accessible birthing models to address vulnerabilities. The model of care and the midwife’s role emerge as pivotal mediators, influencing the overall childbirth experience. While MLUs embody patient-centred, holistic care, the study highlights the potential transferability of these elements to the OUs. In instances where expectations are not met, the identification of elements from the biopsychosocial model within OUs serves as a protective factor, fostering a positive childbirth experience. Key components such as individualised attention, emotional support, respectful and compassionate care emerge as substantial contributors to positive childbirth experiences. These insights provide valuable information for healthcare providers and policymakers, aiming to enhance maternal care practices and foster positive childbirth experiences through the integration of midwifery-led care units in the Spanish Healthcare System. Further research is required to delve deeper into the factors influencing positive birth experiences among women from diverse regions and cultural backgrounds, facilitating more tailored and inclusive approaches to maternal healthcare.

## Data Availability

All data produced in the present study are available upon reasonable request to the authors

