## Supplemental tables for "Beyond the numbers: A phenomenological analysis of women’s childbirth experiences in Spain’s evolving healthcare system"

Table 1. Participants characteristics and birth details.

| Pseudonym | Age range | Parity | Type of birth | Onset of labour | Model of care |
|---|---|---|---|---|---|
| Participant 1 | 30-35 | Primiparous | Instrumental birth | Induced | Combined care |
| Participant 2 | 30-35 | Multiparous | Unplanned caesarean birth | Spontaneous | OU |
| Participant 3 | 40-45 | Multiparous | Spontaneous vaginal birth | Spontaneous | OU |
| Participant 4 | 35-40 | Multiparous | Spontaneous vaginal birth | Spontaneous | OU |
| Participant 5 | 35-40 | Primiparous | Instrumental birth | Induced | OU |
| Participant 6 | 30-35 | Primiparous | Water birth | Spontaneous | MLU |
| Participant 7 | 30-35 | Multiparous | Water birth | Spontaneous | MLU |
| Participant 8 | 30-35 | Primiparous | Spontaneous vaginal birth | Spontaneous | Combined care |
| Participant 9 | 25-30 | Primiparous | Spontaneous vaginal birth | Spontaneous | MLU |
| Participant 10 | 30-35 | Primiparous | Unplanned caesarean birth | Induced | OU |
| Participant 11 | 40-45 | Primiparous | Spontaneous vaginal birth | Spontaneous | MLU |
| Participant 12 | 30-35 | Primiparous | Spontaneous vaginal birth | Spontaneous | Combined care |
| Participant 13 | 25-30 | Multiparous | Spontaneous vaginal birth | Induced | OU |
| Participant 14 | 20-25 | Primiparous | Instrumental birth | Induced | OU |
| Participant 15 | 35-40 | Primiparous | Instrumental birth | Induced | OU |
| Participant 16 | 30-35 | Primiparous | Water birth | Spontaneous | Combined care |
| Participant 17 | 40-45 | Primiparous | Spontaneous vaginal birth | Spontaneous | Combined care |
| Participant 18 | 30-35 | Multiparous | Unplanned caesarean birth | Spontaneous | Combined care |
| Participant 19 | 30-35 | Primiparous | Spontaneous vaginal birth | Spontaneous | MLU |

a- MLU: Midwifery-led unit, aligned with a biopsychosocial model of care.
b- OU: Obstetric unit, aligned with a biomedical model of care.
c- Combined care: transfer from MLU to OU during or after labour.

Table 2. Focus group interview guide.

| |
|---|
| **1. General experience of childbirth** |
| Icebreaker: In one word, how would you describe your childbirth experience? |
| Can you share your childbirth experience and what it meant for you, your baby and your family? |
| **2. Negative aspects of childbirth** |
| What would you change from your childbirth experience? |
| **3. Positive aspects of childbirths** |
| What do you believe is necessary for a childbirth experience to be positive? |
| When prioritising, of all we have discussed, what do you consider was most important during childbirth? |
| **4. Additional aspects** |
| Would you add anything else that has not been mentioned by you or any member of the group? |

Table 3. Themes and sub-themes identified.

| Themes | Subthemes |
|---|---|
| 1. Shaping birth expectations | 1.1 Cultural and social factors |
| | 1.2 Previous obstetric experiences |
| | 1.3 Knowledge, interaction and healthcare accessibility |
| 2. The childbirth essentials | 2.1 One to one care |
| | 2.2 Pain relief strategies |
| | 2.3 Privacy and dignity |
| | 2.4 Respectful care |
| 3. Navigating the “positives” and the “negatives” | 3.1 Meeting birth expectations |
| | 3.2 Meeting elements of “value” in care |
| | 3.3 Midwifery role on “childbirth essentials” and “elements of value” |
